# Epistemic Mistrust and Rigidity mediate the effect of Individual Narcissism on Conspiracy Mentality: Investigating mediators of the narcissism-conspiracy link within the Epistemic Trust Framework

**DOI:** 10.1101/2025.10.05.25337380

**Authors:** Felix Brauner, Nicola-Hans Schwarzer, Peter Fonagy, Chloe Campbell, Julia Griem, Tobias Nolte

## Abstract

**Background:** Prior research has shown that belief in conspiracy theories (CTs) is associated with individual narcissism, encompassing both grandiosity and vulnerability. Initial evidence suggests that this link is mediated by mistrust and suspicion towards others, alongside rigid belief systems such as odd beliefs, fatalism, or delusions.

**Objective:** Within the Epistemic Trust (ET) framework, this study examined whether the narcissism–conspiracy link is accounted for by epistemic stances of mistrust and rigidity formed developmentally.

**Methods:** A cross-sectional design was used, with self-report data from 417 UK-based adults from the non-clinical population. Pathological narcissism, assessed via the Pathological Narcissism Inventory (PNI), was modelled as the independent variable, and conspiracy mentality, assessed with the Conspiracy Mentality Questionnaire (CMQ), as the dependent variable. Epistemic mistrust (Epistemic Trust, Mistrust, and Credulity Questionnaire; ETMCQ) and epistemic rigidity (Epistemic Vice Scale; EVS) were tested as mediators. Structural equation modelling (SEM) with latent variables was conducted.

**Results:** SEM indicated that epistemic mistrust and rigidity fully mediated the effect of narcissism on conspiracy mentality, accounting for 20% of the variance. This mediation pattern held for overall narcissism as well as the grandiosity and vulnerability subdimensions.

**Conclusions:** The influence of subclinical narcissistic traits on conspiracy mentality appears to operate via mistrust and rigidity in epistemic stance. For narcissistic individuals, these may function as defensive strategies to shield their belief systems from external challenge by disengaging from social input. The findings highlight the importance of communication strategies, informed by the ET framework, that promote openness and cognitive flexibility in individuals with heightened narcissism who endorse conspiratorial beliefs.

## Introduction

A “contemporary trend” in psychological research on conspiracy beliefs and their antecedents involves examining pathological personality traits as potential risk factors [1, 2]. Meta-analytic evidence indicates that several such traits—paranoia, schizotypy, psychoticism, delusion-proneness—even in their subclinical expressions, account for meaningful variance in conspiracy belief endorsement [2–4]. Among these, individual narcissism has emerged as particularly influential, with meta-analytic correlations in the range of *r* = .22–.28 [2–4]. Understanding the psychological processes that link narcissism to conspiratorial thinking is thus of growing interest [5]. To date, there has been limited research on the question which epistemic mechanisms play a mediating role in this process. The present study focuses on one such mechanism drawn from the Epistemic Trust (ET) framework: epistemic mistrust and rigidity.

### Theoretical background

#### Effects of individual narcissism on conspiracy beliefs

Recent studies have increasingly explored the influence of so-called “Dark Triad” traits— narcissism, Machiavellianism, and psychopathy—or their extension, the “Dark Tetrad” (including sadism), on belief in conspiracy theories [1, 6]. Accumulating evidence supports the role of these traits in promoting conspiratorial ideation [7–12]. The largest available study, spanning nine countries (n = 13,546), found that individual narcissism significantly predicts conspiracy mentality—a trait-like disposition to endorse a range of CTs—using multi-group SEM [13].

Further meta-analytic results reinforce this, showing that narcissism, conceptualised as beliefs in one’s superiority and sense of entitlement, increases conspiracy beliefs, whereas non-narcissistic self-esteem—positive self-regard characterised by adequacy and satisfaction— reduces it. The effect size for narcissism is substantially greater than that for self-esteem [2–4]. Importantly, large-scale cross-national datasets (n > 50,000) confirm that narcissism remains a significant predictor of conspiracy beliefs even when controlling for other relevant psychological and demographic variables [14, 15].

In short, individual narcissism has been reliably implicated in conspiracy belief endorsement, yet the mediating psychological mechanisms still have to be clearly identified.

#### Effects of narcissistic subdimensions, and mediators of the narcissism-conspiracy link

Nomological network analyses distinguish two interrelated yet distinct subdimensions of pathological narcissism, validated in large-scale studies [16]. Narcissistic grandiosity (NG) reflects traits of superiority, arrogance, exploitativeness, and dominance, along with positive affect linked to self-serving beliefs. In contrast, narcissistic vulnerability (NV) is characterised by defensive self-focus, low self-worth, shame, and emotional reactivity when narcissistic needs are frustrated. Despite these distinctions, both dimensions share a core of antagonistic self-entitlement—marked by manipulation, cynicism, and exploitation—which defines pathological narcissism [16].

Both NG [11, 17] and NV [11, 18] have been shown to predict conspiracy beliefs, even after adjusting for confounders [14]. Although they may operate via partially distinct psychological routes, these effects appear to be at least partly driven by the antagonistic core common to both subtypes [5].

In a review framing conspiracy ideation through self-related motives, Biddlestone et al. [3] proposed that narcissistic individuals may develop a conspiracy mentality as a means of self-protection—preserving a positive self-image and deflecting blame for personal shortcomings. Supporting this view, large-scale, cross-cultural studies (n = 15,000) have shown that heightened conspiracy beliefs among narcissistic individuals are linked to motivational self-enhancement strategies, such as boosting ego through denigrating others (rivalry) or positioning oneself as a heroic figure during crises (heroism) [19]. Nevertheless, it remains unclear which specific *epistemic* mechanisms enable narcissistic individuals to adopt conspiracy beliefs in pursuit of these self-related goals.

Previous research has begun to identify mediators that may account for the narcissism– conspiracy association. Notably, heightened mistrust in others [10, 14] and paranoid ideation [11, 20] strengthen this relationship. Additionally, rigid belief systems—characterised by resistance to contradictory evidence, such as fatalism, odd beliefs, or delusional ideation [10, 11]—and disrupted reasoning styles [21] have been implicated. The link is further amplified in individuals with rigid cognitive reflection styles, who exhibit overconfidence in their reasoning and strong investment in intuitive (rather than rational) beliefs [14].

Taken together, these findings suggest that narcissistic individuals may adopt conspiracy beliefs to protect the self, and that tendencies to mistrust others and to hold on to preconceived beliefs with high rigidity function as mediating mechanisms. However, to date, the question remains open what *epistemic* processes could be related to these mechanisms, i.e., whether specific stances regarding the communication of knowledge may also be relevant in mediating the narcissism-conspiracy-link. The present study addresses this research gap using the Epistemic Trust (ET) framework, which provides a theoretical basis for understanding how epistemic stances shape the transmission and reception of social knowledge. Thus, the distinct potential of applying the ET framework lies in exploring whether specific epistemic stances that distort social knowledge transfer can help explain why narcissistic individuals develop a conspiracy mentality.

#### Investigating the narcissism*–*conspiracy link within the Epistemic Trust framework

The Epistemic Trust (ET) framework conceptualises ET as trust in communicated knowledge—an evolutionarily and developmentally shaped predisposition essential for adaptive social learning [22–24]. High levels of ET reflect openness to the authenticity, relevance, and generalisability of socially transmitted information, fostering psychological resilience over time [22–24]. By contrast, adverse childhood experiences can disrupt ET, producing a hypervigilant stance towards interpersonal communication. Such disruptions manifest as epistemic mistrust and rigidity, potentially limiting access to corrective social information. These maladaptive stances have been linked to pathological personality traits, functional impairments, and enduring psychopathology [25–29].

A series of recent empirical studies has confirmed that disruptions in ET—specifically epistemic mistrust and credulity—are associated with elevated conspiracy mentality [30–35]. However, within the ET framework, Brauner et al. [30] demonstrated that only epistemic mistrust is a robust predictor of conspiracy mentality; the apparent influence of epistemic credulity was fully accounted for by its association with mistrust. This pattern has been replicated in other studies showing that mistrust—but not credulity—retains predictive value when controlling for other variables [32, 33]. Beyond mistrust, epistemic rigidity has also emerged as a significant contributor to conspiracy belief endorsement and susceptibility to misinformation [36, 37]. The associations between ET disruptions and conspiracy beliefs have further been linked to early adversity (e.g., emotional neglect and abuse), impaired personality functioning, and diminished resilience capacities, such as poor mentalizing [32–35, 38].

Building on this growing empirical base, the present study aimed to investigate if the ET framework can help to fill the research gap regarding the narcissism-conspiracy link in terms of how specific *epistemic* stances, distorting social knowledge transfer, may play a mediating role. Drawing on the ET framework, we hypothesised that narcissism is associated with an epistemic stance characterised by, first, mistrust of alternative perspectives due to perceived superiority of one’s own view, and second, rigidity in maintaining and asserting that view within the social environment [39, 40]. Mixed-sample studies that include individuals across the psychopathology spectrum have demonstrated associations between narcissistic traits and disrupted ET [41, 42]. In a large-scale community sample (n = 1,600), Cruciani et al. [43] found that profiles of pathological narcissism—encompassing both grandiose and vulnerable subtypes—are especially associated with higher levels of epistemic mistrust. Experimental findings further suggest that in narcissistic individuals, shame induction via negative evaluation from an authority figure exacerbates state-level epistemic mistrust which points to a self-protecting function of this epistemic stance by turning away from the social environment [44]. These findings support the view that the ET framework offers a viable explanatory model for how specific epistemic stances of mistrust and rigidity may mediate the link between narcissism and conspiracy mentality.

#### The present study

An unresolved question in conspiracy research concerns the epistemic mechanisms through which individual narcissism influences conspiracy ideation. The primary aim of the present study was to investigate whether epistemic mistrust and epistemic rigidity—key constructs from the ET framework—mediate the relationship between narcissistic personality traits (encompassing both grandiosity and vulnerability) and conspiracy mentality, conceptualised as the dispositional tendency underlying belief in specific conspiracy theories. This study extends prior work based on the same dataset, which examined the general impact of ET disruptions on conspiracy mentality [30].

#### Hypotheses

H1: Conspiracy mentality is positively associated with individual narcissism, as well as with narcissistic grandiosity and vulnerability.

H2: Individual narcissism, narcissistic grandiosity, and narcissistic vulnerability are each positively associated with epistemic mistrust and epistemic rigidity.

H3: The relationship between individual narcissism and conspiracy mentality is mediated by epistemic mistrust and epistemic rigidity.

H4: The effects of both narcissistic grandiosity and narcissistic vulnerability on conspiracy mentality are mediated by epistemic mistrust and epistemic rigidity.

## Materials and methods

### Participants

A cross-sectional design was used to recruit a non-clinical UK-based adult sample via Prolific. Data was collected between 29/06/2021 and 21/07/2021. This data was collected as part of a larger parent study “Probing Social Exchanges” and was given REC Approval by Research Ethics Committee of Wales 3 (reference: 12/WA/0283) and HRA Approval by NHS (IRAS project ID: 103075). Participants were monetarily compensated (£7.50/hour) and gave their written informed consent. The data collection was fully anonymised and did not contain any information which could identify individual participants. Although Prolific’s sampling algorithm aimed to capture a socioeconomically diverse population, the final sample showed some biases: younger adults were overrepresented (mean age = 33.38 years, SD = 11.11), women comprised 68% of the sample, and levels of educational attainment were relatively high (37% with higher education and 21% with postgraduate qualifications). A detailed description of the sample can be looked up at Table 1.

**Table 1.** Sociodemographic characteristics of the sample (n=417)

| Sample Characteristics | N | % |
| --- | --- | --- |
| Gender |  |  |
| Male | 134 | 32.1 |
| Female | 282 | 67.6 |
| Other | 1 | 0.2 |
| Education |  |  |
| No qualifications | 4 | 1.0 |
| Other qualifications (not listed) | 5 | 1.2 |
| Vocational level 1 (GCSE < 5 A*-C) | 24 | 5.8 |
| Vocational level 2 (GCSE > 5 A*-C) | 23 | 5.5 |
| A level (vocational level 3) | 119 | 28.5 |
| Higher education (or equivalent) | 155 | 37.2 |
| Postgraduate education (or equivalent) | 87 | 20.9 |
| Household income |  |  |
| Less than £10,000 | 134 | 32.1 |
| £10,000-£20,000 | 99 | 23.7 |
| £20,000-£35,000 | 114 | 27.3 |
| £35,000-£50,000 | 42 | 10.1 |
| £50,000-£75,000 | 22 | 5.5 |
| £75,000-£100,000 | 3 | 0.7 |
| Not stated | 2 | 0.5 |

*Note.* N = 417. Participants were on average 33.38 years old (SD = 11.11).

### Variables and Instruments

#### Individual narcissism

Individual narcissism was assessed using the Pathological Narcissism Inventory (PNI), a 52-item measure capturing both narcissistic grandiosity and narcissistic vulnerability [45]. Items are rated on a 5-point Likert scale (1 = “strongly disagree” to 5 = “strongly agree”). The validated higher-order structure of the PNI was employed [45]. The grandiosity factor (NG) comprises the subscales Exploitativeness, Grandiose Fantasy, and Self-Sacrificing Self-Enhancement, while the vulnerability factor (NV) includes Contingent Self-Esteem, Devaluing, Entitlement Rage, and Hiding the Self. Taken together, a total score for pathological narcissism (PN) was computed across all subscales. The scale demonstrated excellent internal consistency in the current sample (PN α = .96; NG α = .91; NV α = .94). A confirmatory factor analysis (CFA) using the seven z-standardised subscales as manifest indicators confirmed the latent factor structure, with satisfactory fit indices (χ²(7, 405) = 16.155, *p* = .024; RMSEA = .057 [.020–.094]; CFI = .994; SRMR = .019).

#### Conspiracy mentality

Conspiracy mentality was assessed using the Conspiracy Mentality Questionnaire (CMQ), a five-item self-report measure capturing the general tendency to endorse conspiracy theories [46]. Items reflect broad suspicions about hidden power structures (e.g., “I think that there are secret organizations that greatly influence political decisions”) and are rated on a scale from 0% (“certainly not”) to 100% (“certainly”). The CMQ has demonstrated sound psychometric properties, including convergent and discriminant validity, normal distribution, test–retest reliability, and cross-cultural applicability, supporting its interpretation as a latent trait underlying specific conspiracy beliefs [46]. In the present study, the CMQ showed good internal consistency (α = .84). A confirmatory factor analysis (CFA) using the five z-standardised items as manifest indicators confirmed the latent factor structure (χ²(3, 405) = 8.525, *p* = .036; RMSEA = .068 [.015–.123]; CFI = .994; SRMR = .018).

#### Epistemic mistrust

Epistemic mistrust was measured using the Mistrust subscale of the Epistemic Trust, Mistrust, and Credulity Questionnaire (ETMCQ). The ETMCQ [27] is a 15-item self-report instrument assessing openness to, or disruption of, social knowledge transmission. Responses are given on a 7-point Likert scale (1 = “strongly disagree” to 7 = “strongly agree”). The Mistrust subscale (ETMCQ-EM) comprises five items (e.g., “I don’t usually act on advice that I get from others even when I think it’s probably sound”), capturing a stance of pervasive scepticism toward interpersonal communication and a reluctance to be influenced by others. Validation studies have supported the scale’s reliability and cross-cultural robustness [27, 29, 47–50]. In our sample, internal consistency was acceptable (α = .70), and CFA confirmed the latent structure, with all five z-standardised items loading significantly (χ²(5, 405) = 5.078, *p* = .406; RMSEA = .006 [.000–.070]; CFI = 1.000; SRMR = .020).

#### Epistemic rigidity

Epistemic rigidity was assessed via the Rigidity subscale of the Epistemic Vice Scale (EVS), which conceptualises epistemic vices as dispositional barriers to the acquisition and exchange of knowledge [36]. The Rigidity subscale (EVS-ER) consists of six items (e.g., “I tend to feel sure about my views even if I don’t have much evidence”), rated on a 5-point scale (1 = “strongly disagree” to 5 = “strongly agree”). This subscale captures a tendency toward inflexible adherence to one’s beliefs irrespective of disconfirming evidence. The EVS has demonstrated strong construct validity and favourable psychometric properties [36, 37]. In the current study, internal consistency for the Rigidity subscale was satisfactory (α = .74), and CFA supported the latent factor structure (χ²(7, 405) = 11.584, *p* = .115; RMSEA = .040 [.000–.080]; CFI = .990; SRMR = .027).

### Statistical data analysis

All analyses were conducted using IBM SPSS Statistics 29 and IBM SPSS AMOS 29. The dataset contained less than 3% missing values. Little’s MCAR test indicated that these were missing completely at random (*p* > .05). Further inspection revealed 12 participants with incomplete questionnaire data due to a technical issue at the beginning of the survey. Given the minimal proportion of missing data and negligible impact on statistical power, these cases were excluded via listwise deletion [51], resulting in a final sample of 405 participants.

Descriptive statistics were computed for all variables and compared to values reported in the respective validation studies. Pearson’s correlation analyses were conducted to examine bivariate associations, with Bonferroni corrections applied to adjust the alpha level for multiple comparisons. As Mardia’s test revealed violations of multivariate normality, structural equation modelling (SEM) was performed using maximum likelihood estimation with robust standard errors and 10,000 bootstrap samples.

SEM was used to test hypothesised pathways with “conspiracy mentality” as the endogenous variable. In the first model, “individual narcissism” was entered as the exogenous variable. In a second step, “narcissistic grandiosity” and “narcissistic vulnerability” were tested as separate exogenous variables in distinct models to avoid multicollinearity (r = .68 between the two subscales) and to maintain comparability with previous studies that adopt this modelling approach [10, 14, 20, 21]. In all models, “epistemic mistrust” and “epistemic rigidity” were entered as mediators.

Model fit was evaluated using multiple indices (52): χ², the root mean square error of approximation (RMSEA) with 90% confidence interval, the comparative fit index (CFI), and the standardised root mean square residual (SRMR). Excellent fit was defined by a non-significant χ², RMSEA ≤ .06, CFI ≥ .95, and SRMR ≤ .06; acceptable fit was indicated by RMSEA ≤ .08, CFI ≥ .90, and SRMR ≤ .08. Given the large sample size, a significant χ² was anticipated [53]. Direct and indirect effects were estimated using bootstrap confidence intervals derived from 10,000 bootstrap samples.

## Results

### Descriptive characteristics of the data

We first examined the descriptive characteristics of all key variables in relation to values reported in prior validation studies. Mean scores for pathological narcissism were M = 3.03 (PNI total), M = 3.17 (NG), and M = 2.90 (NV) (see Table 2), which fall within the expected range for non-clinical populations but are somewhat elevated relative to normative scores reported in the PNI validation study (PNI total: M = 2.38; NG: M = 2.62; NV: M = 2.20; [45]). This elevation may reflect the composition of our sample, which overrepresented younger, highly educated individuals—a demographic shown to report slightly elevated narcissistic traits [54].

**Table 2.**
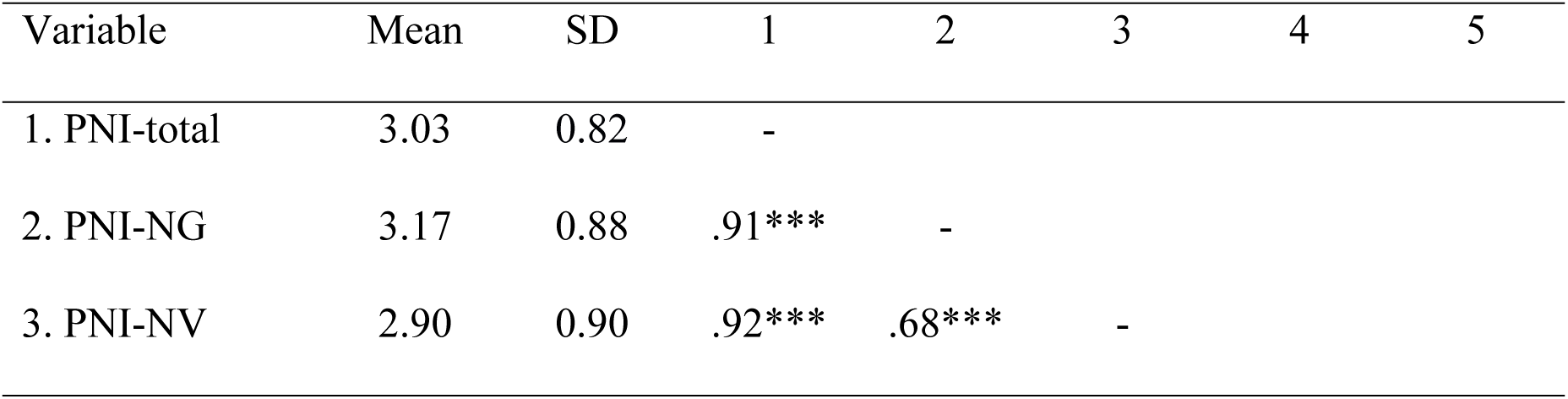
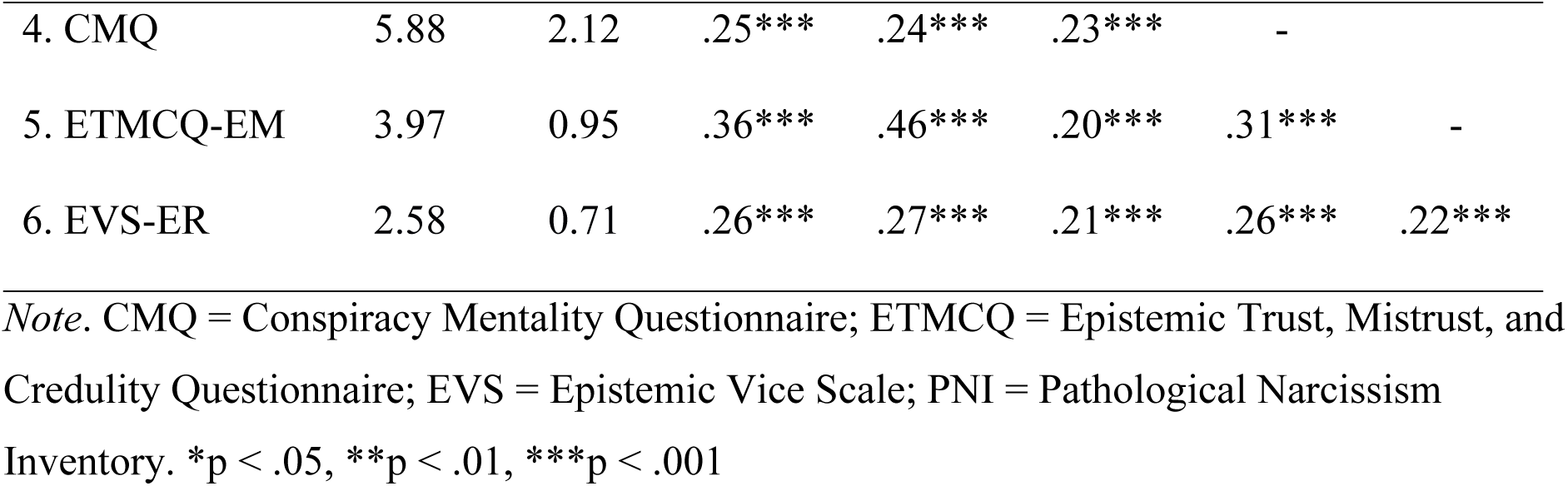
Descriptive statistics and intercorrelations for all study variables.

| Variable | Mean | SD | 1 | 2 | 3 | 4 | 5 |
| --- | --- | --- | --- | --- | --- | --- | --- |
| 1. PNI-total | 3.03 | 0.82 | - |  |  |  |  |
| 2. PNI-NG | 3.17 | 0.88 | .91*** | - |  |  |  |
| 3. PNI-NV | 2.90 | 0.90 | .92*** | .68*** | - |  |  |
| 4. CMQ | 5.88 | 2.12 | .25*** | .24*** | .23*** | - |  |
| 5. ETMCQ-EM | 3.97 | 0.95 | .36*** | .46*** | .20*** | .31*** | - |
| 6. EVS-ER | 2.58 | 0.71 | .26*** | .27*** | .21*** | .26*** | .22*** |

*Note.* CMQ = Conspiracy Mentality Questionnaire; ETMCQ = Epistemic Trust, Mistrust, and Credulity Questionnaire; EVS = Epistemic Vice Scale; PNI = Pathological Narcissism Inventory. \* $p < .05$ , \*\* $p < .01$ , \*\*\* $p < .001$

For conspiracy mentality, the mean score of M = 5.88 was slightly below the M = 6.30 observed in the UK CMQ validation study [46], suggesting that while participants showed moderate agreement with conspiracy beliefs, they did not exhibit high levels of ideological commitment—consistent with the general trend in non-clinical samples [2–4]. Mean scores for epistemic mistrust (M = 3.97) and epistemic rigidity (M = 2.58) were in line with those reported in validation studies [27, 36].

### Correlation across variables

Bivariate correlations are reported in Table 2. Narcissistic grandiosity (NG) and narcissistic vulnerability (NV) were strongly correlated (r = .68, *p* < .001). All narcissism variables showed significant associations with conspiracy mentality: total PN (r = .25, *p* < .001), NG (r = .24, *p* < .001), and NV (r = .23, *p* < .001). Regarding the mediators, epistemic mistrust (EM) correlated significantly with PN (r = .36, *p* < .001), NG (r = .46, *p* < .001), and NV (r = .20, *p* < .001). Epistemic rigidity (ER) was also significantly associated with PN (r = .26, *p* < .001), NG (r = .27, *p* < .001), and NV (r = .21, *p* < .001). Conspiracy mentality (CM) correlated significantly with EM (r = .31, *p* < .001) and ER (r = .26, *p* < .001), and the two mediators were significantly intercorrelated (r = .22, *p* < .001).

### Structural equation modelling

Fig. 1 presents the SEM results with total narcissism as the independent variable. The model demonstrated good fit: χ²(214, 405) = 489.429, *p* < .001; RMSEA = .056 [.050–.063]; CFI = .922; SRMR = .060. Bootstrap analysis (10,000 samples) confirmed significant paths from pathological narcissism to epistemic mistrust (β = .52 [.43–.61], *p* < .001) and epistemic rigidity (β = .36 [.26–.46], *p* < .001).

**Fig. 1.**
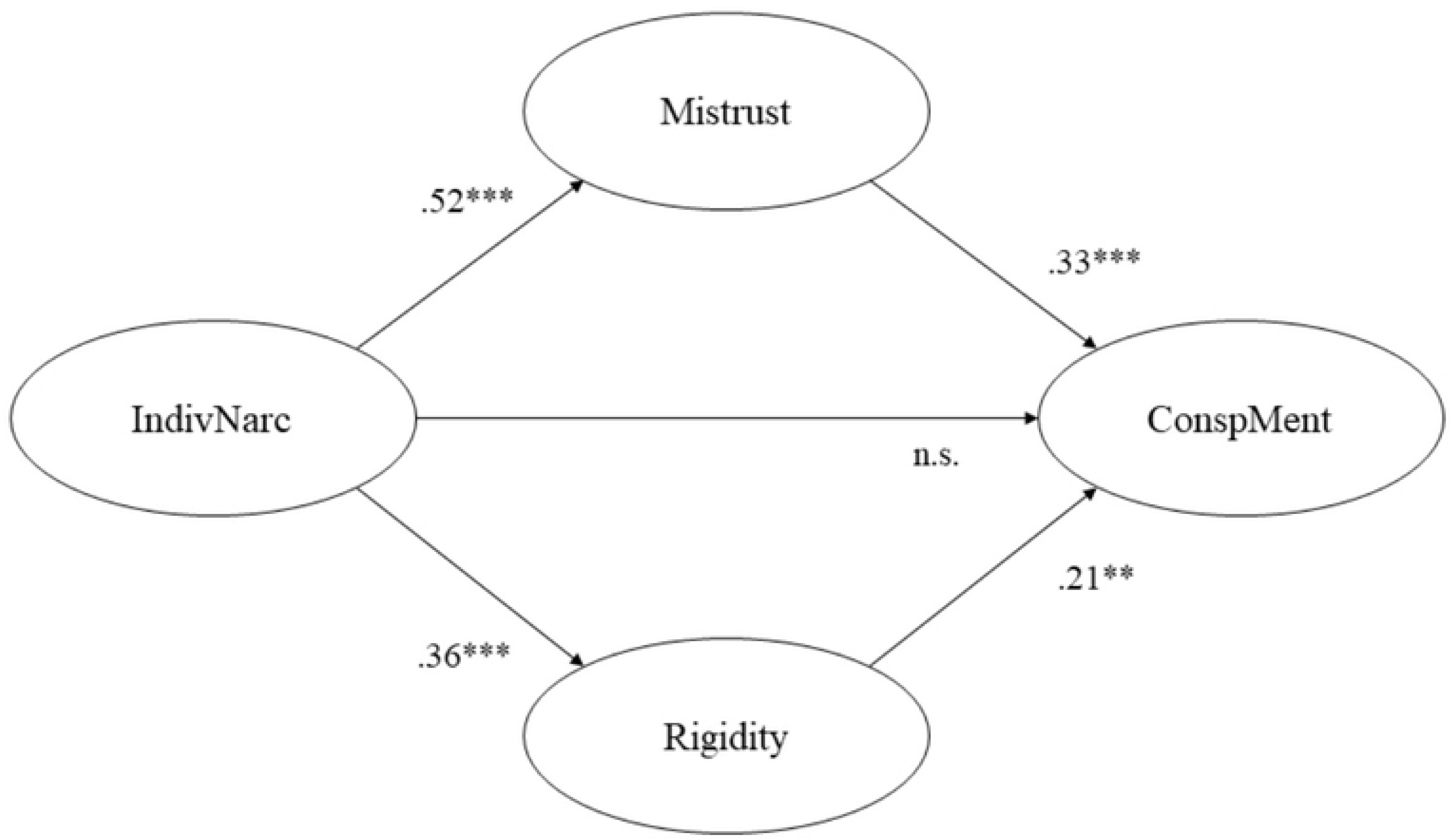
Epistemic mistrust and rigidity mediating the relationship between individual narcissism (PNI-total) and conspiracy mentality (CMQ)

There was no significant direct effect of pathological narcissism on conspiracy mentality (β = .03 [–.10–.14], *p* = .730) when accounting for the mediators. Both epistemic mistrust (β = .33 [.20–.46], *p* < .001) and epistemic rigidity (β = .21 [.09–.33], *p* < .01) were independently associated with conspiracy mentality. Indirect effects confirmed full mediation of the narcissism–conspiracy link via epistemic mistrust (β = .17 [.09–.28], *p* < .001) and epistemic rigidity (β = .08 [.02–.15], *p* < .01). The total effect of pathological narcissism on conspiracy mentality remained significant (β = .28 [.18–.36], *p* < .001), with the model explaining 19.6% of the variance in conspiracy mentality.

### Follow-up models: narcissistic grandiosity and vulnerability

To examine the unique effects of narcissistic subdimensions, we tested two separate SEM models due to the high correlation between grandiosity and vulnerability (r = .68), which raised concerns about multicollinearity—evidenced by standardised beta coefficients exceeding +1.00 when both were entered simultaneously. Both models demonstrated good fit: grandiosity model: χ²(143, 405) = 278.857, *p* < .001, RMSEA = .048 [.040–.057], CFI = .936, SRMR = .063; vulnerability model: χ²(161, 405) = 359.830, *p* < .001, RMSEA = .055 [.048–.063], CFI = .930, SRMR = .058.

Fig. 2 and Fig. 3 present the SEM results with the narcissistic subdimensions of grandiosity and vulnerability as independent variables. Using 10,000 bootstrap samples, narcissistic grandiosity was significantly associated with epistemic mistrust (β = .31 [.21–.42], *p* < .001) and epistemic rigidity (β = .26 [.15–.37], *p* < .01). Narcissistic vulnerability also showed significant positive associations with epistemic mistrust (β = .56 [.46–.64], *p* < .001) and epistemic rigidity (β = .37 [.27–.48], *p* < .001). However, neither grandiosity (β = .10 [–.01–.21], *p* = .123) nor vulnerability (β = –.02 [–.15–.11], *p* = .820) exhibited direct effects on conspiracy mentality once epistemic stances were included.

**Fig. 2.**
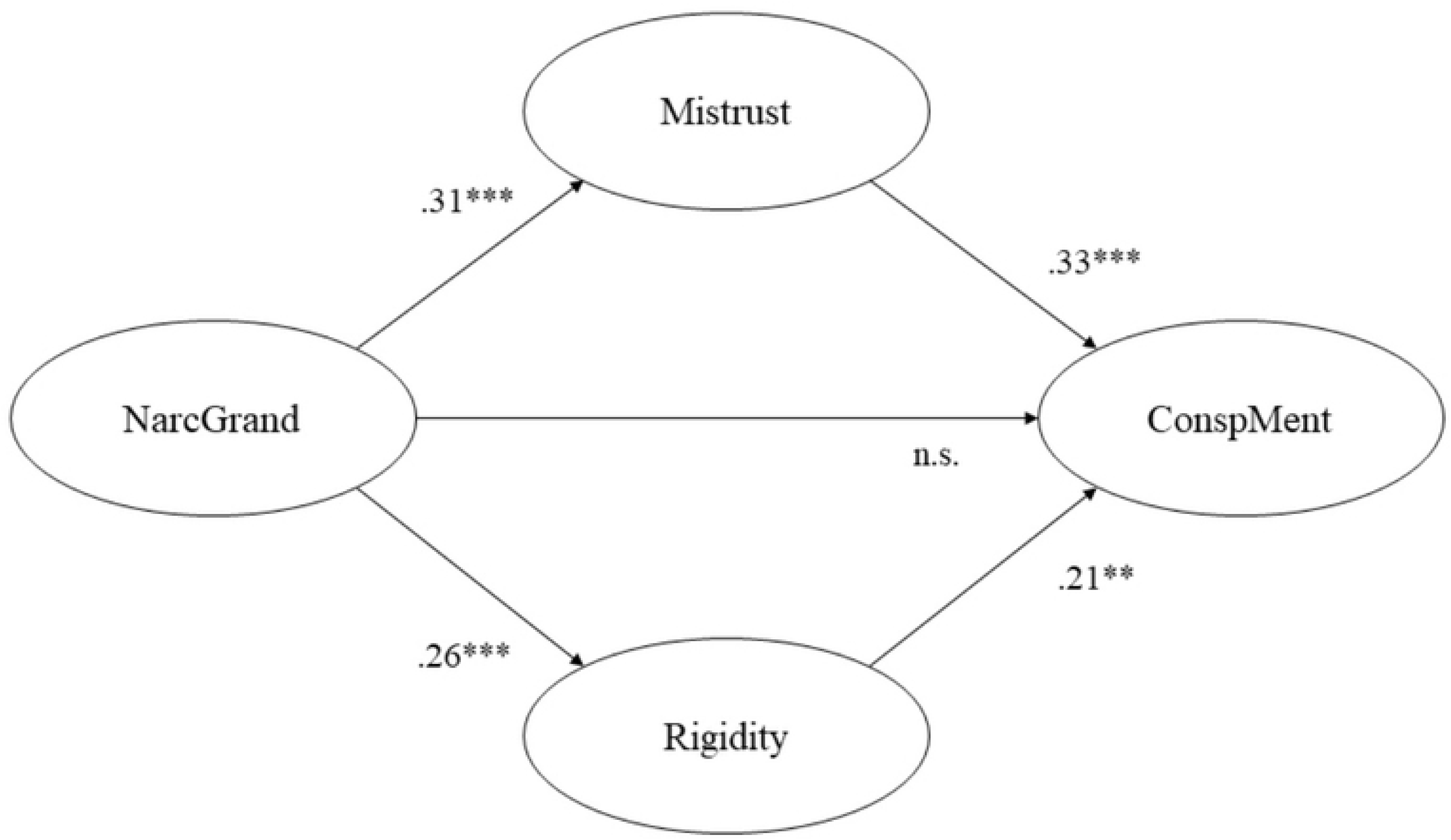
Epistemic mistrust and rigidity mediating the relationship between narcissistic grandiosity (PNI-NG) and conspiracy mentality (CMQ)

**Fig. 3.**
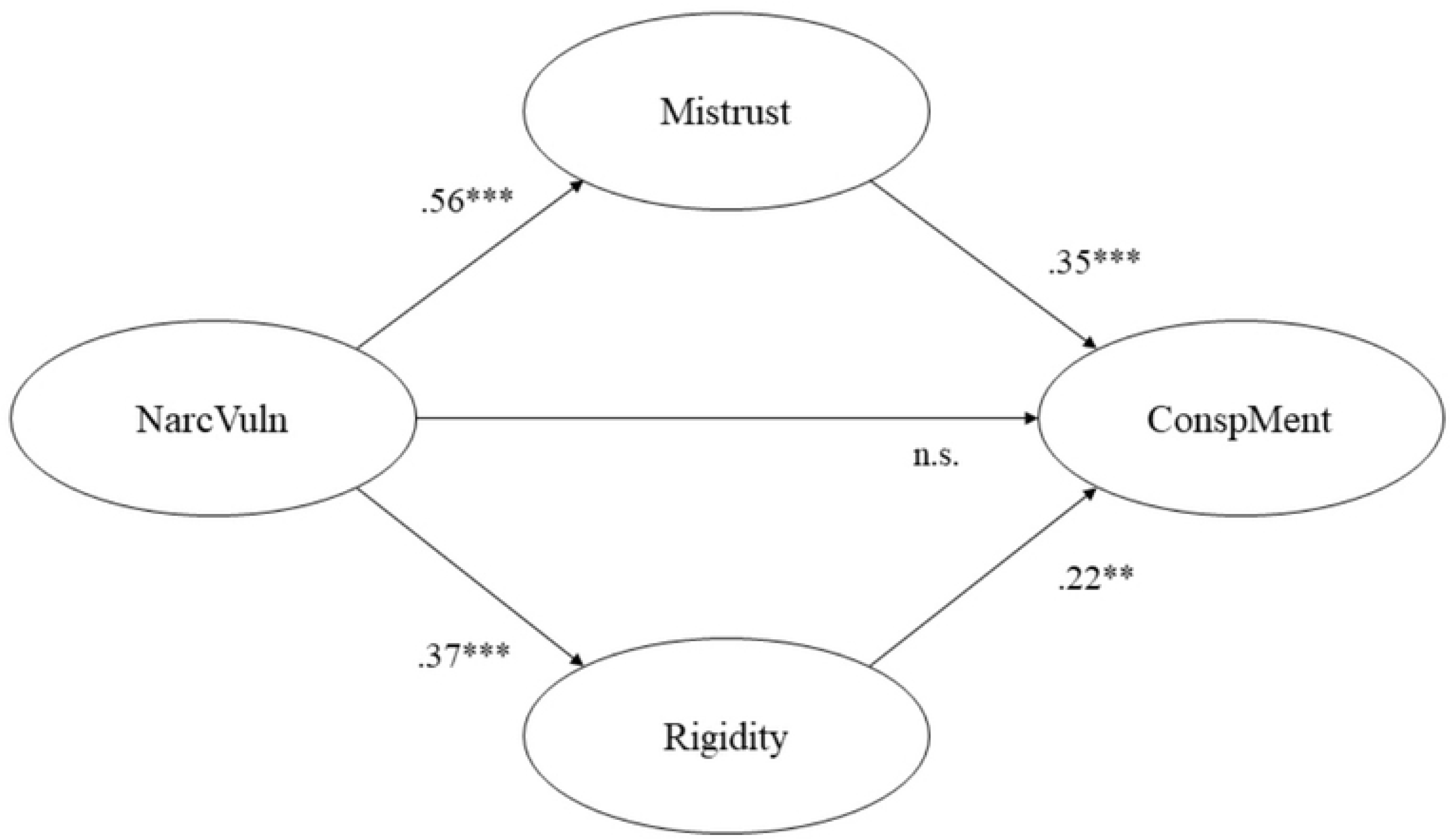
Epistemic mistrust and rigidity mediating the relationship between narcissistic vulnerability (PNI-NV) and conspiracy mentality (CMQ)

In both models, epistemic mistrust and rigidity were directly associated with conspiracy mentality: for grandiosity, EM (β = .33 [.21–.44], *p* < .001), ER (β = .21 [.09–.32], *p* < .01); for vulnerability, EM (β = .35 [.22–.48], *p* < .001), ER (β = .22 [.10–.34], *p* < .01). The indirect paths from narcissistic grandiosity to conspiracy mentality were significant via epistemic mistrust (β = .10 [.04–.18], *p* < .01) and rigidity (β = .05 [.01–.12], *p* < .05). Similarly, the effects of narcissistic vulnerability were fully mediated by epistemic mistrust (β = .20 [.10–.31], *p* < .001) and rigidity (β = .08 [.03–.16], *p* < .05).

Total effects were significant for both narcissistic grandiosity (β = .26 [.15–.36], *p* < .001) and vulnerability (β = .26 [.16–.35], *p* < .001), explaining 20.2% and 19.6% of the variance in conspiracy mentality, respectively.

## Discussion

Guided by the Epistemic Trust (ET) framework, this study explored whether epistemic mistrust and epistemic rigidity mediate the relationship between individual narcissism and conspiracy mentality. Drawing on cross-sectional data from a UK-based online sample, and using structural equation modelling, we found that both epistemic mistrust and rigidity fully mediated the associations between pathological narcissism—including both grandiosity and vulnerability— and conspiracy mentality.

These findings expand on prior work using the same dataset, which demonstrated that epistemic mistrust plays a central role in conspiracy ideation, even when accounting for other psychological predictors [30].

In our correlation analyses, conspiracy mentality was significantly associated with overall individual narcissism (r = .25), as well as with its two subdimensions—narcissistic grandiosity (r = .24) and narcissistic vulnerability (r = .23)—thus supporting H1. These effect sizes align with recent meta-analytic estimates, which report moderate associations (r = .22–.28) between narcissistic traits and conspiracy beliefs [2–4], and with findings demonstrating relations of both grandiosity and vulnerability with conspiracy beliefs [5, 14, 17, 18].

We further found significant associations between individual narcissism and both epistemic mistrust (r = .36) and epistemic rigidity (r = .26), with each subdimension—grandiosity and vulnerability—also showing positive correlations with both epistemic variables (see Table 2), thereby confirming H2. Our findings corroborate previous evidence showing that narcissistic traits are associated with ET disruptions [41–44]. In line with expectations, conspiracy mentality was also significantly associated with both epistemic mistrust (r = .31) and epistemic rigidity (r = .26), echoing findings from other ET-informed studies [30–35].

Using structural equation modelling, we showed that the relationship between narcissistic traits—both grandiosity and vulnerability—and conspiracy mentality was fully mediated by epistemic mistrust and rigidity, thereby confirming H3 and H4. While earlier studies explored various mediators amplifying the influence of narcissism on conspiracy beliefs [e.g., 10, 11, 14, 20, 21], our study is the first to directly test *epistemic* stances as potential mediators. Our mediation models including both epistemic mistrust and rigidity as mediators accounted for 20% of the variance in conspiracy mentality, a magnitude comparable to that observed in prior research examining other mediators such as paranoia [20], odd beliefs and fatalism [10], or cognitive reflection and thinking styles [14, 21]. Taken together, our findings contribute to a growing body of evidence clarifying the psychological mechanisms through which narcissism shapes conspiracy ideation [5]. At the same time, the substantial unexplained variance in our models indicates that additional mediators should be explored in future research.

By its grounding in the Epistemic Trust (ET) framework, our study reveals a broader understanding of the narcissism-conspiracy-link that goes beyond previous research. Based on the ET framework, high levels of epistemic mistrust and rigidity emerge in early interpersonal environments as strategies to cope with a communication perceived as untrustworthy, which is closely related to the development of pathological personality components, such as pathological narcissism. While these adaptations may serve short-term protective functions, they hinder openness to social learning in more benign contexts later in life [24–26]. Pathological narcissism, from this perspective, is characterised by a perceived superiority and entitlement that coexists with deep mistrust of others (viewed as inferior) and rigid adherence to one’s own viewpoint (considered superior) which is corroborated by studies on associations between narcissistic traits and ET disruptions [41–44]. In narcissistic individuals, these epistemic stances of mistrust and rigidity disrupt the transmission of socially relevant knowledge by impairing their reciprocal communicative engagement [39, 40]. Following this line of research, our findings suggest that the heightened tendency to believe in conspiracy believes of narcissistic individuals can be explained by the fact that their development of pathological narcissistic traits is accompanied by epistemic stances of mistrust and rigidity, which impairs their social knowledge transfer.

Applying the ET framework also provides a new perspective on models framing conspiracy ideation as a self-related coping strategy [3]. Biddlestone et al. [3] proposed that conspiracy beliefs allow narcissistic individuals to preserve a favourable self-image and deflect blame for personal failures. Similarly, large-scale cross-cultural studies confirm that narcissistic self-protection strategies—such as denigrating others or adopting heroic narratives—predict higher conspiracy belief [19]. Our findings are broadly consistent with these accounts, but allow the additional assumption that specific *epistemic* motives—such as the desire to shield oneself from disconfirming information—may underlie these self-protective strategies. Specifically, epistemic mistrust and rigidity may serve to close off engagement with the social world, thereby reducing the likelihood of external challenge and preserving an idealised self-concept [39–44].

It is important to note that, although our study was situated within the Epistemic Trust (ET) framework, we did not include the stance of epistemic credulity. Preliminary studies examining the relationship between epistemic disruptions and conspiracy beliefs have found significant associations with credulity, as measured by the ETMCQ [30–35]. However, when controlling for relevant confounding factors such as authoritarianism, loneliness, attachment insecurity [30], paranoid distress and psychiatric symptoms [32], or need for closure and general psychopathology [33], only epistemic mistrust remained a significant predictor of conspiracy mentality. Indeed, Brauner et al. [30] showed that the apparent effect of credulity is fully accounted for by its shared variance with mistrust. For these reasons, we chose to focus on mistrust (rather than credulity) in our SEM models. Moreover, our preliminary analyses indicated that including credulity as an additional mediator would have compromised model stability (tolerance < .01; VIF > 10). Nevertheless, future studies with larger samples and more statistical power should further explore the potential role of credulity—particularly its interplay with mistrust and rigidity—in shaping the relationship between narcissism and conspiracy beliefs.

### Implications for future research

Our findings contribute to growing evidence that ET disruptions—particularly mistrust and rigidity—can explain how pathological personality traits, such as individual narcissism, increase susceptibility to conspiracy mentality. Previous research has shown that these epistemic disruptions are associated with other predictors of disordered personality functioning, including low levels of reflective function and impaired personality structure [34, 55, 56]. There is also initial evidence suggesting that other Dark Tetrad traits (e.g., Machiavellianism, psychopathy, sadism) may operate through similar pathways as narcissism—especially via rigid cognitive styles, closed-mindedness, and mistrust in interpersonal and institutional communication [6]. Thus, future research could extend the ET framework by examining whether epistemic rigidity and mistrust also mediate the effects of other Dark Tetradtraits on conspiracy mentality.

Beyond conspiracy beliefs, epistemic disruptions have been linked to broader forms of misinformation susceptibility. For instance, individuals high in mistrust and rigidity are more likely to reject public health guidance and to endorse politically motivated fake news [31, 33, 35]. Recent research has also demonstrated that a worldview combining narcissism, conspiracy mentality, mistrust, and rigidity increases vulnerability to misinformation [57. 58], and promotes its active spread—particularly via social media [59]. Accordingly, future research should examine whether the same mechanisms we identified—epistemic mistrust and rigidity—also predict susceptibility to, and dissemination of, misinformation in domains beyond conspiracy belief.

Previous research has shown that narcissistic individuals exhibit heightened sensitivity to social evaluation and rejection, responding with increased stress reactivity and vigilance [60]. In line with this, experimental research grounded in the ET framework has demonstrated that shame induction—triggered via negative performance feedback—reduces epistemic trust and increases epistemic mistrust in narcissistic individuals [44]. These findings support the core assumption of the ET framework that, while epistemic stance is conceptualised as a trait-like disposition, its expression is context-dependent [61]. In particular, social situations that induce shame—such as criticism of personal beliefs or worldviews—may reinforce mistrust and rigidity in individuals high in narcissistic traits, thereby amplifying conspiracy mentality. Future experimental studies could explore this hypothesis further by investigating whether situationally induced social stress (e.g., via the Trier Social Stress Test, TSST) leads to increased conspiracy belief through transient disruptions in epistemic stance, as measured by implicit tasks such as the Epistemic Trust, Mistrust, and Credulity Implicit Task (ETMCIT).

A further avenue for research concerns the developmental roots of these dynamics. According to the ET framework, pathological personality traits—including narcissism—are shaped by early adverse interpersonal experiences, which contribute to the formation of rigid and mistrustful epistemic stances [23–26]. Meta-analytic evidence supports this view, indicating significant associations between adverse childhood experiences and both narcissistic grandiosity and vulnerability [62]. Childhood adversity has also been linked to general and specific conspiracy beliefs, often mediated by reduced interpersonal trust and cognitive flexibility [63–65]. The ET framework offers a plausible explanatory pathway: preliminary findings suggest that the impact of early adversity on conspiracy mentality is mediated by epistemic mistrust and rigidity [34, 35]. Within this context, reduced mentalizing—a core component of the ET framework—may also play a mediating role, given its demonstrated association with conspiracy thinking [32, 33, 55]. Future studies may explore if interrelationships with childhood adversity or mentalizing deficits – in addition to epistemic disruptions – also have a strengthening influence on the narcissism-conspiracy link.

### Limitations

Several limitations should be considered when interpreting our findings.

First, the study relied on a self-selected online sample from the UK, which introduces potential selection bias. Although Prolific’s sampling methods aimed for socio-demographic diversity, our sample overrepresented younger adults (M = 33 years) and women (68%). These characteristics are known to influence both narcissism [54] and conspiracy mentality [8], and may also affect the strength of associations between the two—typically being more pronounced in samples including older individuals and more men [2]. Replication in more demographically representative samples is therefore warranted.

Second, while we used the Pathological Narcissism Inventory (PNI), which captures narcissistic grandiosity and vulnerability and is widely validated [16], emerging research has drawn attention to the Trifurcated Model of Narcissism (TriMN). This model emphasises a three-factor structure comprising antagonism (core to both grandiosity and vulnerability), agentic extraversion (specific to grandiosity), and narcissistic neuroticism (specific to vulnerability) [16]. Notably, recent studies suggest that only antagonism predicts conspiracy beliefs [66]. Although the PNI loads strongly on antagonism [54], future studies could benefit from directly assessing TriMN dimensions using more differentiated instruments [67].

Third, the cross-sectional design of our study prevents inferences about directionality over time. While prior longitudinal findings support the pathway from narcissism to conspiracy belief rather than the reverse [68], future research should test our mediation model longitudinally to confirm the hypothesised direction: narcissism → mistrust/rigidity → conspiracy mentality.

Fourth, our study was conducted in the UK, a country with comparatively low levels of conspiracy endorsement [69]. In contexts with higher baseline levels of conspiracy beliefs— such as states characterised by authoritarianism and corruption [46]—the influence of personality variables like narcissism appears attenuated [12]. Future research could explore how epistemic mistrust and rigidity interact with cultural context by replicating this study in such environments.

Finally, our findings emerged from a WEIRD (Western, Educated, Industrialised, Rich, Democratic) cultural context [70]. Meta-analyses have shown that the narcissism–conspiracy association is stronger in WEIRD samples than in non-WEIRD ones [71]. Replicating this study in non-WEIRD settings is necessary to determine whether the mediating effects of epistemic stances generalise across cultural contexts.

In sum, while our study offers novel insights into the relationship between individual narcissism, epistemic mistrust and rigidity, and conspiracy mentality, these findings should be interpreted with the above limitations in mind.

## Conclusion

Our findings show that the effect of individual narcissism—including both grandiosity and vulnerability—on conspiracy mentality is fully mediated by epistemic mistrust and epistemic rigidity. By applying the Epistemic Trust (ET) framework, our study offers a novel explanation for the well-established narcissism-conspiracy link by showing *epistemic* mechanisms as mediators for the first time. The study reveals that distorted communication of knowledge, related to the epistemic stances of mistrust and rigidity, is crucial why narcissistic individuals develop a conspiracy mentality.

In general, this model may help to explain why individuals high in narcissistic traits have been described as a “hard-to-reach group” in conspiracy belief research [4]. Rather than being simply resistant to alternative views, they may be actively defending against the perceived threat of social influence. As such, interventions aimed at directly correcting their conspiracy beliefs through fact-checking or confrontation are likely to provoke resistance. Our findings suggest that effective intervention strategies should instead aim to address the underlying epistemic stance characterised by mistrust and rigidity. Thus, the integration of the ET framework may offer a more viable path toward engagement and belief revision in this group.

## Abbreviations

CT: conspiracy theory
ET: epistemic trust
NG: narcissistic grandiosity
NV: narcissistic vulnerability

## Ethics approval and consent to participate

REC Approval was given by Research Ethics Committee of Wales 3 (reference: 12/WA/0283) and HRA Approval was given by NHS (IRAS project ID: 103075). Participants gave their written informed consent to participate and received financial compensation (at a rate of £7.50 per hour) for participating via the online survey platform Prolific.

## Availability of data and materials

the data underlying the results presented in the study are available from the Research Ethics Committee of Wales 3 (contact via) for researchers who meet the criteria for access to confidential data.

## Author contributions

FB and TN conceptualised the study design, in consultation with JG and NS; FB drafted the first version of the manuscript, reviewed by TN; TN coordinated the data collection, and JG performed the data curation; FB computed the descriptives and correlational analyses, and NS computed the CFA and SEM analyses; FB created the figures and tables; all authors critically revised the manuscript and approved the final version to be published.

## Acknowledgements

No acknowledgements to be declared.

